# Spatial neglect therapy with the augmented reality app “Negami” for active exploration training; A randomized controlled trial

**DOI:** 10.1101/2023.03.26.23287770

**Authors:** Britta Stammler, Kathrin Flammer, Thomas Schuster, Marian Lambert, Oliver Neumann, Michael Lux, Tamara Matuz, Hans-Otto Karnath

## Abstract

**Background:** A widely applied and effective rehabilitation method in stroke patients suffering from spatial neglect is the ‘visual exploration training’. Patients improve their ipsilesional bias of attention and orientation by training of exploration movements and search strategies towards the contralesional side of space. Here we investigate the effectiveness of the augmented reality (AR)-based app *“Negami”* for the treatment of spatial neglect in a randomized control trial. *Negami* combines a visual exploration training with active, contralesionally oriented rotation of eyes, head, and trunk.

**Methods:** Twenty patients with spatial neglect were randomly assigned to the experimental *Negami* group or to a group receiving standard neglect therapy. Over a period of two weeks, both groups received five training sessions per week (à 25 minutes). Neglect behavior was assessed weekly over a five-week period, with the *Negami* therapy group receiving a second follow-up assessment at one-to-two-month intervals after completion of training.

**Results:** Both groups improved significantly. While the *Negami* therapy group improved in four of five neglect tests used, the standard therapy group improved in only one of these tests. We observed significantly better improvement in the *Negami* group already after the first week of training. This difference was also significant after the end of the training as well as one week after the end of training and remained stable one to two months after the end of treatment.

**Conclusion:** *Negami* can be used as an effective alternative or addition to current standard neglect therapy, and may even be superior to it.

**Trial Registration** at “OSF Preregistration”, Registration DOI: https://doi.org/10.17605/OSF.IO/H6XN5 and at WHO approved public trial registry “German Clinical Trials Register”, DRKS-ID: DRKS00031446

## Introduction

Spatial neglect is the dominant cognitive disorder following right hemispheric brain damage in humans ^1,2^. Typically, it is evoked by strokes in the right middle cerebral artery region that damage the perisylvian network, consisting of the superior/middle temporal, parietal, and ventrolateral frontal cortex. Such patients act as though the left side of space has vanished. The patient’s eyes and head are sustainedly oriented to the side of the brain lesion, which is typically the right side ^3-5^. The patient’s visual and tactile exploration activity is shifted to the right side; information located on the left side is disregarded. Thus, many approaches to treating spatial neglect focus on performing exercises and tasks that stimulate patients to actively orient toward the side that is being neglected, such as, e.g., visual searching and picture description tasks, or reading and copying tasks ^6-10^. This therapeutic approach, also known as “visual exploration training” or “visual scanning training,” aims to increase exploration movements and compensating search techniques, which shall improve neglect behavior in everyday scenarios.

The visual exploration training seems to be particularly effective when the active eye and head movement is combined with an active rotation of the trunk in this direction ^11^. Wiart et al. (1997) found significant improvements in neglect symptoms with both acute and chronic neglect who received this combined exploration therapy compared to controls. Their finding was in line with studies that had observed that proprioceptive stimulation by trunk rotation or the vibration of posterior neck muscles reduce spatial neglect and thus may have an additional effect on the efficacy of neglect treatment ^12-14^. The augmented reality (AR)-based app *Negami* represents an attractive new tool that builds on these findings ^15^. It is based on the principle that patients are playfully motivated to orient themselves to their neglected side of the real room by (a) following and (b) searching for a virtual element (an origami bird), actively exploring space by turning their eyes, head, and trunk. *Negami* uses the principle of AR in which the visual, real world is augmented by a virtual figure via a video camera of an electronic device such as a tablet.

There is an increasingly growing interest in augmenting existing neuropsychological treatment methods with apps. This can be explained by two things: First, the desire to maximize the effectiveness of treatment, and second, the ability to continue treatment after the patient is discharged. The aim of the present study was to investigate the effectiveness of using the app *Negami* for the treatment of spatial neglect. It should be compared with the use of standard treatment of spatial neglect in three different rehabilitation facilities. Our goal was to enrich the established visual exploration trainings by an innovative new offer, encouraging patients in a playful way to actively explore space by turning their eyes, head, and trunk.

## Material and Methods

### Participants

Twenty patients with right-sided stroke and spatial neglect participated in the study. They were recruited from three different rehabilitation facilities (Kliniken Schmieder, Stuttgart-Gerlingen; Neurological Rehabilitationcenter Quellenhof, Bad Wildbad; Kreiskliniken Reutlingen, Reutlingen) and were randomly assigned to either the experimental *Negami* therapy group or the standard neglect therapy group (n=10 each). Demographic and clinical details are given in Table 1. Structural imaging was acquired by computed tomography (CT) or magnetic resonance imaging (MRI) instead as part of the clinical routine procedure carried out for all stroke patients in the acute phase at stroke-onset (Fig. 1). For MRI, we used the FLAIR scans. Lesion maps were normalised into 1 × 1 × 1 mm^3^ MNI space using SPM (https://www.fil.ion.ucl.ac.uk/spm) and the Clinical Toolbox ^16^. Patients with tumors or patients in whom scans revealed no obvious lesions were not included. All participants gave their informed consent to participate in the study, which was conducted in accordance with the ethical standards of the 1964 Declaration of Helsinki and was approved by the Ethical Committee at the Medical Faculty of Tübingen University (373/2021B02).

**Table 1.** Demographic and clinical data of all 20 right brain damaged neglect patients.

|  | *Negami* therapy group | Standard neglect therapy group |
| --- | --- | --- |
| Number | 10 | 10 |
| Sex (M/F) | 8/2 | 6/4 |
| Age (years) | 61.3 (15.2) | 60.7 (12.45) |
| Etiology | 5 infarcts, 5 hemorrhages | 4 infarcts, 6 hemorrhages |
| Post-stroke interval (days) | 138.4 (192.1) | 84 (33.86) |
| Hemiparesis (% present) | 100 | 100 |
| Letters cancellation test (CoC) | 0.42 (0.28) | 0.49 (0.29) |
| Bells test (CoC) | 0.38 (0.29) | 0.34 (0.3) |
| Copying task (N omitted) | 4.4 (1.51) | 4.3 (1.64) |
| Line bisection (EWB) | 0.33 (2.5) | 0.23 (0.17) |

Data are presented as mean (SD); CoC, Centre of Cancellation <sup>17</sup>; EWB, Endpoint weightings bias (McIntosh et al., 2017).

**Figure 1.**
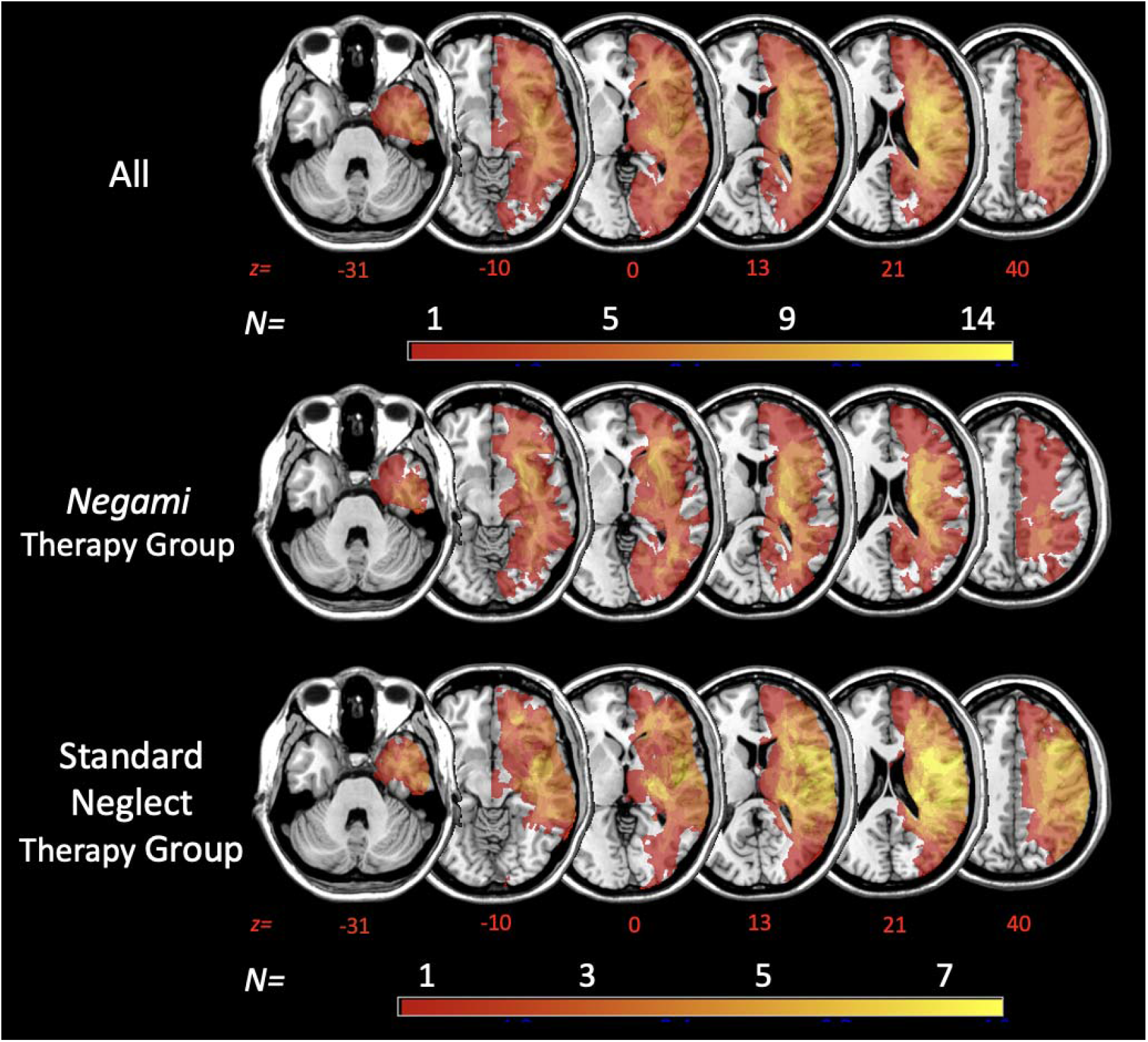
Simple overlay plots. Overlaps of normalized acute lesions are shown for all patients (*N* = 20) and for patients of the *Negami* Therapy Group (*N* = 10) and the Standard Neglect Therapy Group (*N* = 10) on the ch2-template in MNI space via MRIcron ^18^. Displayed axial slices refer to z-coordinates -31, -10, 0, 12, 21 and 40 mm. The color of the voxels represents the number of patients with damage to this voxel (*Nmin* = 1; *Nmax* = 14 or *Nmax* = 7, respectively).

The inclusion criterion for this study was the presence of spatial neglect. In addition to clinical behavioral observations, diagnostic criteria had to be met in at least two of the following four neglect tests: The Letter Cancellation Test ^19^, the Bells Test ^20^, a Copying Task ^21^, and a Line Bisection Task ^22^. All four neglect tests were performed on a Samsung S7+ tablet with screen dimensions 285×185mm. The severity of spatial neglect in the cancellation tasks was determined by calculating the center of gravity of the target stimuli marked in the search fields, i.e. the Center of Cancellation (CoC; ^17^). A CoC value ≥ 0.08 indicated left-sided spatial neglect ^17^. The Copying Task consisted of a complex scene consisting of four objects (fence, car, house, tree), points were assigned based on missing details or whole objects. One point was given for a missing detail, two for a whole object. The maximum number of points is therefore eight. A score higher than 1 (i.e. > 12.5% omissions) indicated spatial neglect ^21^. In the Line Bisection Task ^22^, patients were presented with four different line lengths eight times each, i.e. 32 lines in total. The cut-off value for spatial neglect was an ‘endpoint weightings bias (EWB)’ value ≥ 0.07 ^23^.

### The “Negami” App

The “*Negami*” App was developed for use on a tablet but could also be used on a mobile phone ^15^. For the present experiment, we used an Apple iPad Pro 12.9” 3rd generation. The app allowed to add (augment) a virtual element (origami bird) to the video stream produced by the camera of the tablet. More detailed information on the technical implementation and design of the app can be found in Stammler et al. (2023). The “Negami” app provides two different tasks (Tasks A and B, see below) that each participant performed in succession while using arm movements in combination with trunk rotations. An example patient performing the *Negami* tasks can be seen here: https://youtu.be/fyZ_PVWljp4 or in Stammler et al. (2023) at Multimedia Appendix 2.

#### Task A: “Follow the bird”

The first task of the patient is to follow the virtual origami bird through real space. The patient’s straight-ahead eye/head/body orientation is set as ‘0’. Beginning at set point ‘0’, the bird flies with sinusoidal movements towards one side of space (in neglect patients towards the neglected, contralesional side). While doing so, the patient sees an orange circle in the center of the screen (Fig. 2). The bird should be held in this circle while performing the task. As soon as the patient fails to successfully follow the bird and keep it in the circle during its flight, the patient is provided with an additional orientation aid: a blue compass needle appears showing the patient in which direction the bird is located (see Fig. 2A). The task is considered successfully completed as soon as the bird has finished its trajectory and the bird has been positioned centrally in the orange circle. During the performance the patient receives auditory feedback; every time the patient manages to get the bird into the orange circle, a short, bright tone is presented. If the patient manages to keep the bird continuously in the orange circle, the tone is presented every two seconds. When the task has been successfully completed, the patient receives feedback auditorily again through a different tone and visually through the change of the color of the circle to green.

**Figure 2.**
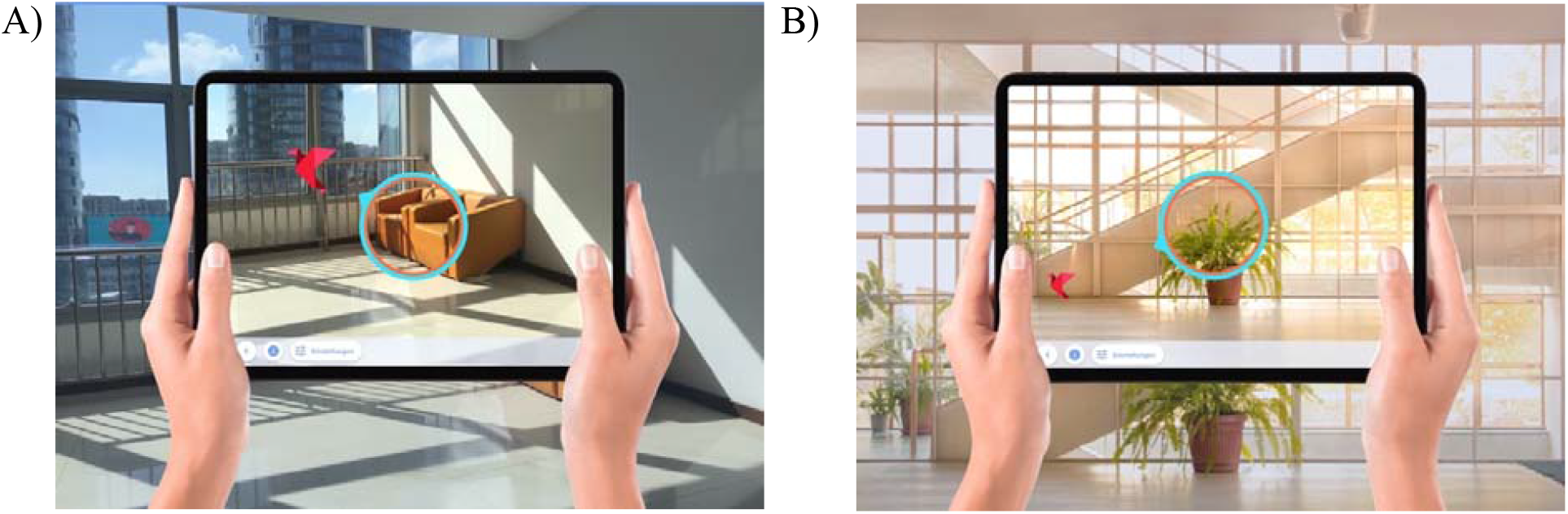
**(A)** Task A “Follow the bird”. The patient has the task to follow the flying origami bird and to keep the bird within the orange/blue circle. **(B)** Task B “Find the bird”. The patient has to search for the bird that has been hidden by the therapist somewhere in the surrounding room (here: at the corner located at the foot of the stairs) and has to transfer it into the orange/blue circle.

With the end of a successful trial, the trail can be repeated or the difficulty level can be adjusted to easy, medium, or difficult (cf. Tab. 1). The difficulty levels are saved as templates.

**Table 1a.**
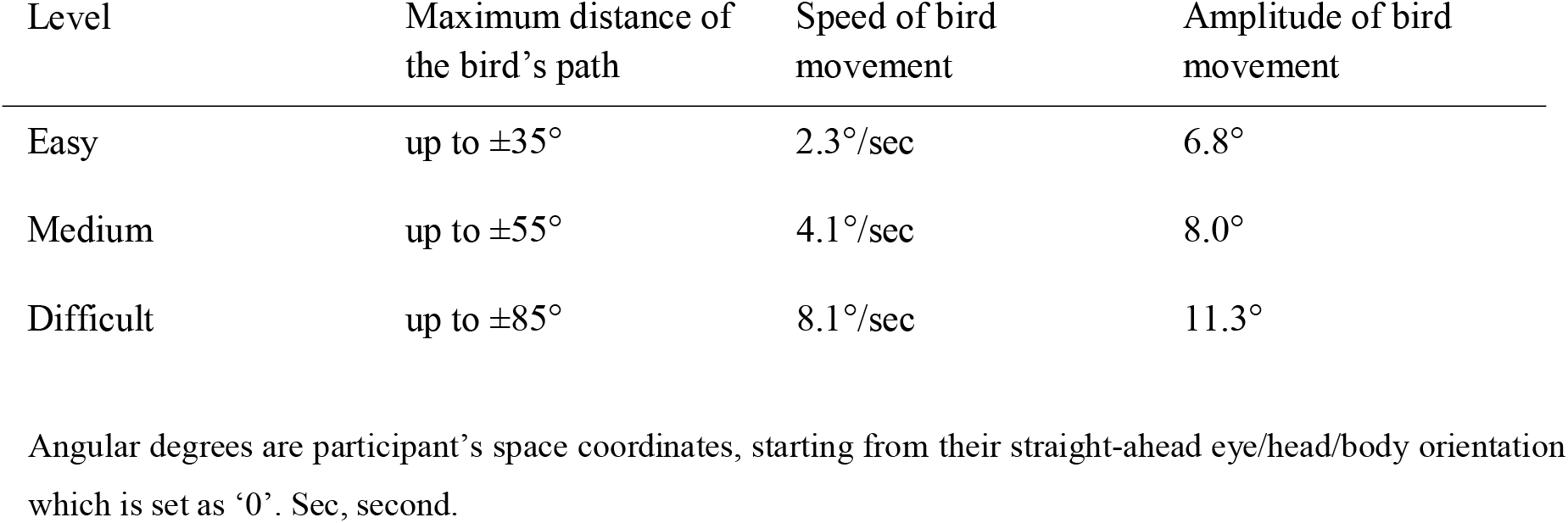
Difficulty levels provided with Task A “Follow the bird”.

#### Task B: “Find the bird”

In the second task of the “Negami” app, the therapist hides the virtual origami bird in the room surrounding the patient. For this purpose, the patient’s straight-ahead eye/head/body orientation again is set as ‘0’. Starting from this location, the therapist hides the bird somewhere on the left/right side of the patient without the patient seeing it. The area in which the bird can be hidden by the therapist is predefined by the app, depending on the chosen difficulty level of the task (for more details see ^15)^. Once the therapist has positioned the bird in space, the patient is then instructed to find the bird and, once found, to position it centrally into the orange circle (cf. Fig. 2B). Should the patient show difficulties in finding the bird, it is possible to provide the patient with orientation assistance by turning on the blue compass needle. The task is successfully solved when the patient has found the bird and placed it in the center of the circle. According to Task A, the circle then turns green and the patient hears a bright tone signaling the successfully solved task. After successful completion the task can be repeated or changed in difficulty (cf. Tab. 2)

**Table 2.** Difficulty levels provided with Task B “Find the bird”.

| Level | Extension of the area (along the horizontal dimension of the surroundings) within which the bird can be hidden |
| --- | --- |
| Easy | 0° to -40° [or 0° to +40°] |
| Medium | -40° to -75° [or +40° to +75°] |
| Difficult | -75° to -90° [or +75° to +90°] |

Angular degrees are participant’s space coordinates, starting from their straight-ahead eye/head/body orientation which is set as ‘0’. Negative values indicate positions to the participant’s left, while positive values indicate positions to the participant’s right.

### Procedure

#### Intervention

Over a period of 2 weeks, the neglect patients of the *Negami* therapy group received five training sessions per week, using the *Negami* app. Each session lasted about 25 minutes. They completed Task A in the first half and Task B in the second half of each training session. The first training session always started with the lowest difficulty level. If the difficulty level was increased in the course of the training, the training session on the following day always started with the training level at which the training was stopped the day before. The criterion for success in advancing to the next difficulty level was that the task was successfully completed three times in a row. The return to a lower difficulty level was indicated when the task was not solved two times in a row. In order to avoid patient frustration when the tasks were not solved, Task A was automatically terminated after 30 seconds if the bird completed its trajectory and the patient did not manage to transfer it into the circle. For Task B, the task was automatically terminated after 90 seconds. The termination was scored as an unsolved task. The neglect patients of the standard neglect therapy group received the standard treatment for neglect of the respective rehabilitation institution. This consisted in all facilities of five training sessions per week, performing a standard visual scanning training and exploration therapy. Each session lasted about 25 minutes.

#### Diagnostic examinations

Before and after intervention both groups were examined five times overall (cf. Fig. 3), using the four diagnostic neglect tests (Letter Cancellation, Bells Test, Copying Task, Line Bisection Task) described above. In addition, a fifth test was performed. Similar to *Negami* Taks B (see above), the patient was instructed to find the hidden bird. However, in this test condition (termed ‘Exploration Test’) no bird was hidden but exploratory movements of the patient were recorded, allowing to calculate mean dwell time of the corresponding viewing angle degree of the space surrounding the neglect patient.

**Figure 3.**
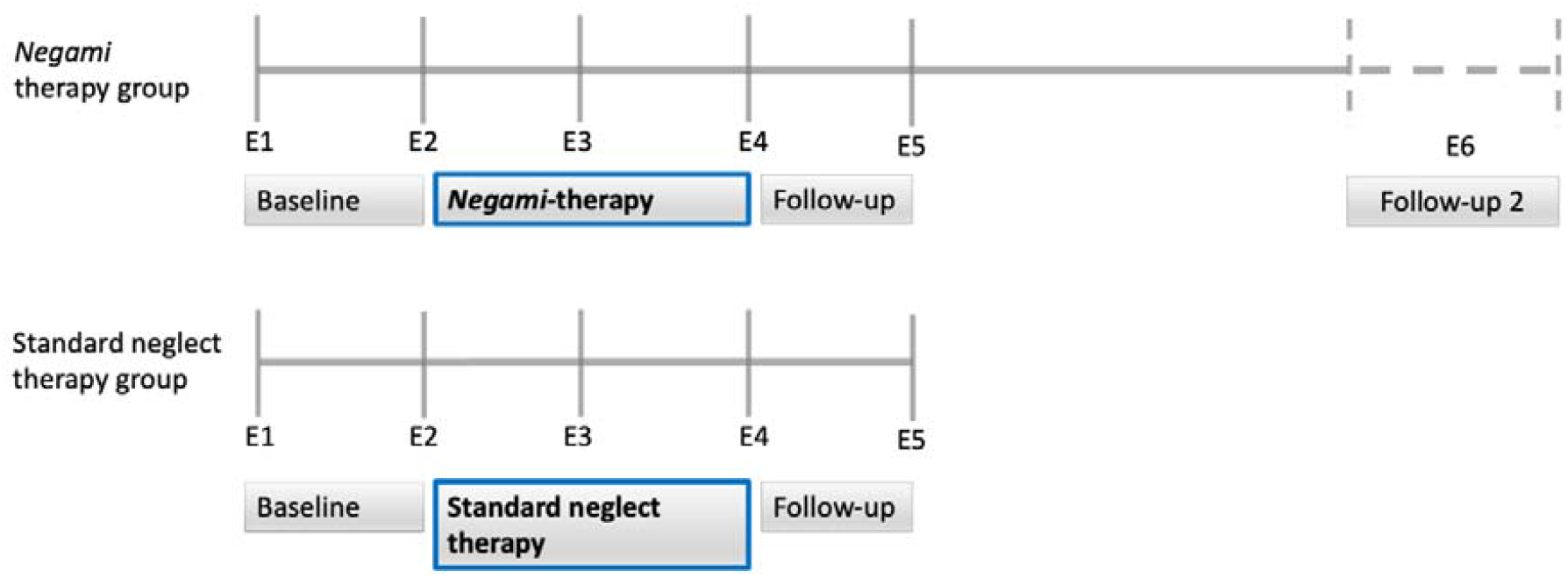
Experimental design. Five diagnostic examinations (E1 to E5) were performed weekly in both the *Negami* therapy group and the standard neglect therapy group. Additionally, the *Negami* group was examined at a time interval of one to two months after completion of training.

Two of the five diagnostic examinations were performed before the start of intervention, allowing to control for spontaneous recovery (Fig. 3; E1 and E2). After the first week of training, a third examination was performed (Fig. 3; E3). After the end of the intervention, the patients of both groups were examined two further times: immediately after and one week after the end of the training (Fig. 3; E4 and E5). Additionally, the *Negami* therapy group was examined at a time interval of one to two months after completion of training (Fig. 3; E6).

## Results

### Descriptive results of the training with Negami

Patients from the *Negami* therapy group completed most days of the two-week training period at the highest level of difficulty (cf. Fig. 4). With the last training session 8 of 10 patients from the *Negami* group reached the difficulty level “difficult” in Task A; two of 10 patients reached the level “medium”. Within the two-week training period, three of the patients from the *Negami* group had to return once to a lower difficulty level because they did not solve Task A at a certain level of difficulty two times in a row. On average, for Task A, difficulty level “medium” was reached after 2.8 (SD=0.92) training days and the difficulty level “difficult” after 4.9 (SD=2.2) days.

**Figure 4.**
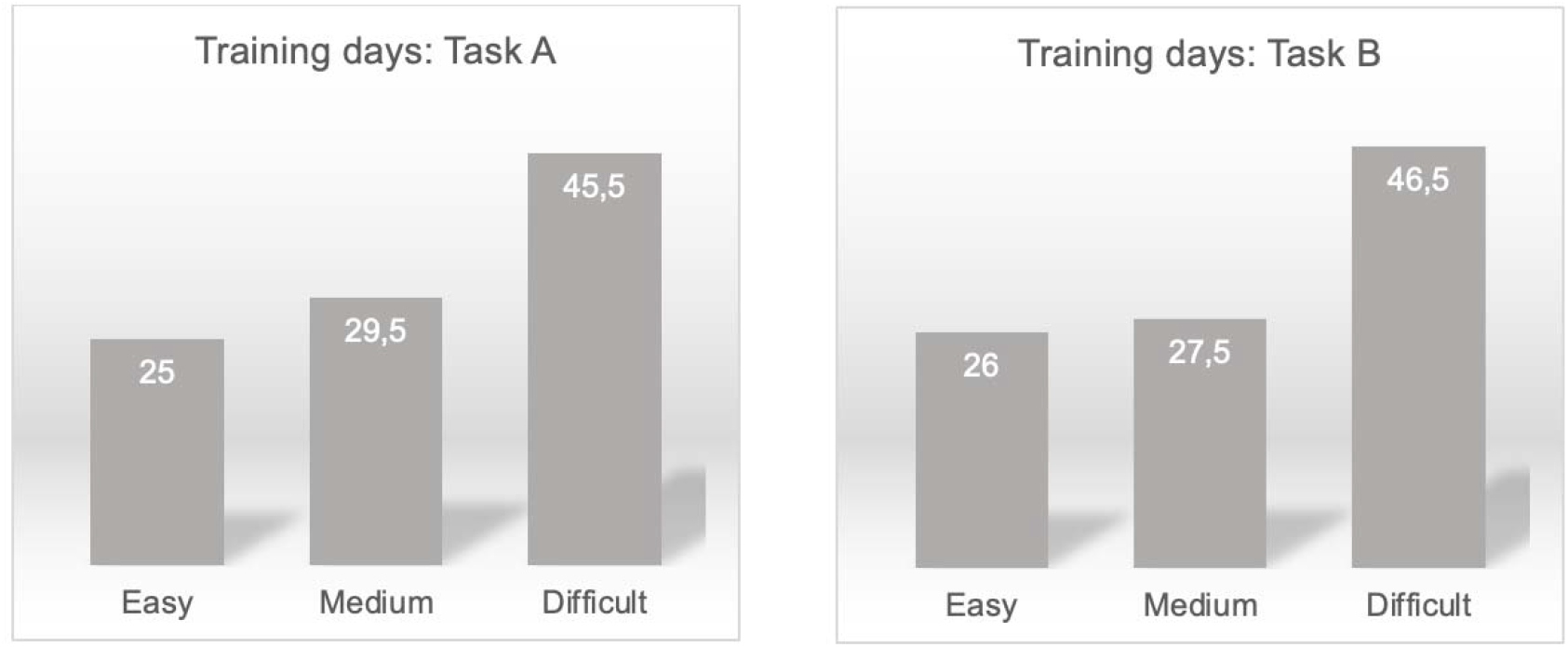
Absolute number of training days that ended with the three difficulty levels ‘easy, medium, difficult’ of Tasks A and B of the *Negami* app. Since each of the 10 patients received 10 training sessions, the maximum number is 100 days.

In Task B, 9 of the 10 patients from the *Negami* group reached the level “difficult” with the last training session; one of 10 patients made it to the level “medium”. None of the patients had to return to a lower difficulty level during the period of therapy. On average, the difficulty level “medium” in Task B was reached after 3.3 (SD=1.49) training days and the difficulty level “difficult” after 6.2 (SD=2.2) days.

### Control for spontaneous recovery

Figure 5 gives an overview on the results of the *Negami* therapy group and the group with standard neglect therapy over all diagnostic examinations. In both groups, we found no significant differences between measurement time points E1 and E2 for all five neglect tests (*Negami* therapy group: dependent t-tests for all five neglect tests: *p*>0.26; standard neglect therapy group: for all five neglect tests *p*>0.23). This indicates that spontaneous remission could be excluded in both groups when treatment started. For the following analyses, we thus combined the two measurement time points E1 and E2 into one (‘baseline’) variable, separate for each of the five diagnostic neglect tests.

**Figure 5.**
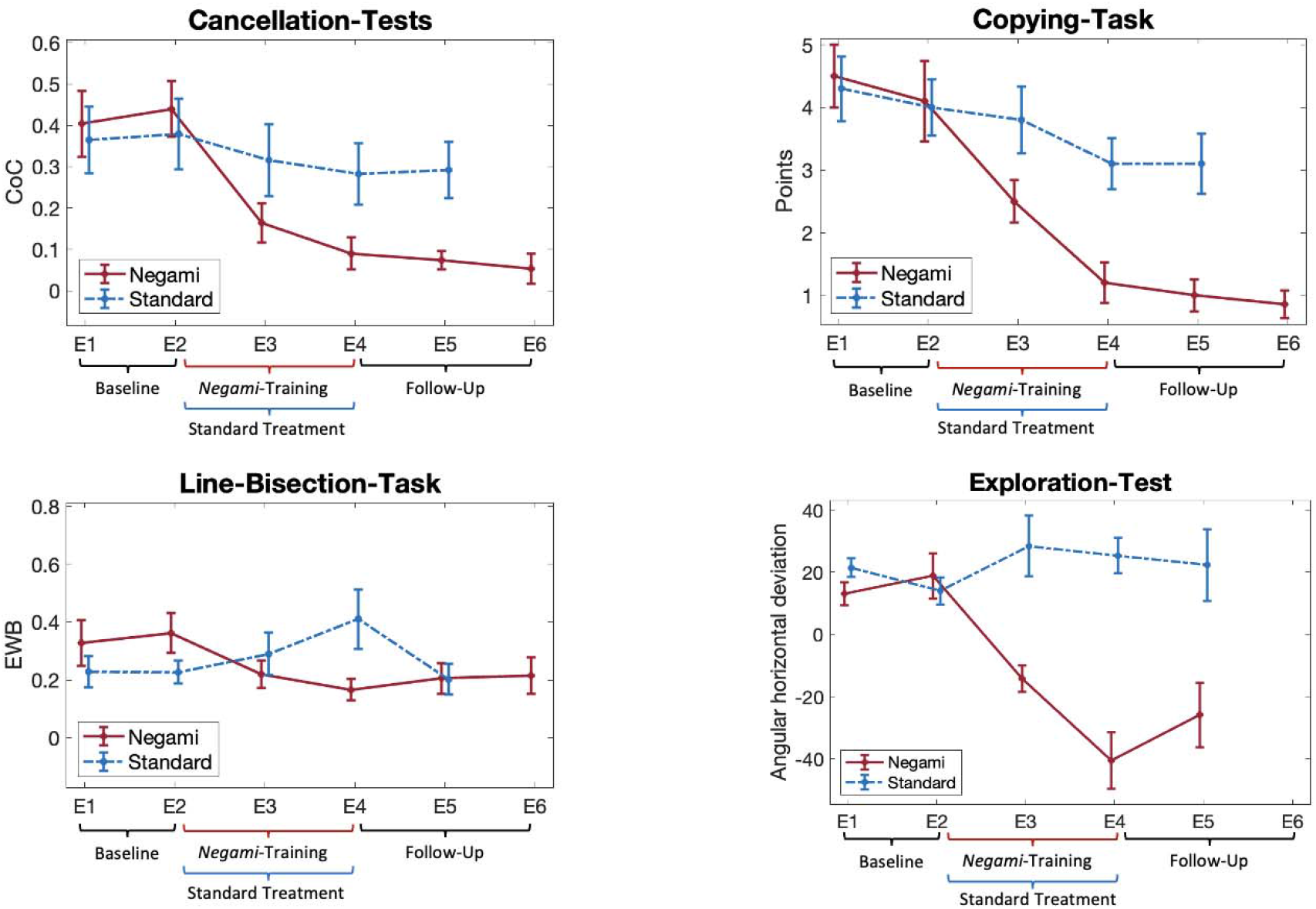
Results in the Cancellation-Tests (performance in the Letter- and the Bells-Cancellation-Tests have been averaged for the sake of clarity), Copying-Task, Line-Bisection-Task, and the ‘Exploration Test’ for the *Negami* therapy group (*red*) and the group with standard neglect therapy (*blue*) over the different diagnostic examination time points (E1 to E6).

### Treatment effects

We conducted ANOVAs of repeated measures with factors group (*Negami* therapy vs. standard neglect therapy) for diagnostic examination time points (baseline, E3, E4, E5), using IBM SPSS Statistics (Vers. 28; ^24^). Statistical significance was defined as *p* < 0.05. Significant interaction effects were found in all five neglect tests (Letter-Cancellation-Test: F(3,54)=2.38, *p*=0.11, η^2^=0.18; Bells-Cancellation-Test: with Greenhouse-Geisser correction F(3,36)=6,49, *p*<0.01, η^2^=0.31; Copying-Task: F(3,54)=8.3, *p*=<0.01, η^2^=0.32; Line-Bisection-Task: with Greenhouse-Geisser correction F(3,31)=3.4, *p*=0.03, η^2^=0.17; ‘Exploration Test’: F(3,39)=5.35, *p*=0.03, η^2^=0.29).

For post-hoc analyses, pairwise t-tests were conducted to examine the within-subject factors with a Bonferroni-corrected significance level of *p* < 0.0125. For the *Negami* therapy group, pairwise t-test comparisons for ‘baseline’ versus time point E4, i.e. the examination right after termination of the intervention, revealed significant differences in four of five diagnostic neglect tests (Letter-Cancellation-Test: t(9)=4.95, *p*<0.01, *d*=0.2; Bells-Cancellation-Test: t(9)=5.7, *p*<0.01, *d*=0.19; Copying-Task: t(9)=6.02, *p*<0.01, *d*=1.63; Line-Bisection-Task: t(8)=3.75, *p*=0.01, *d*=0.2). A clear numerical, but no significant difference was found for the ‘Exploration Test’ (t(6)=4.47, *p*=0.02). For the group with standard neglect therapy, comparisons for ‘baseline’ versus time point E4 revealed one significant improvement in the Copying-Task (t(9)=2.4, *p*=0.01, *d*=1.38). No significant differences were observed for the other four examination tests (Letter-Cancellation-Test: t(9)=1.88, *p*=0.05; Bells-Cancellation-Test: t(9)=0.98, *p*=0.35; Line-Bisection-Task: t(9)=-1.92, *p*=0.04; ‘Exploration Test’: t(8)=-1.49, *p*=0.09).

To examine if there was any significant change in neglect behavior between the first and the second week of training, i.e. between E3 and E4, further paired t-tests were performed. In the *Negami* therapy group only the ‘Exploration Test’ revealed a significant difference (t(6)=3.18, *p*=0.01; *d*=21.88; all other four diagnostic examination tests: *p*>0.02). In the standard neglect therapy group, the only significant difference between E3 and E4 was found for the Copying-Task (t(9)=2.69, *p*=0.01, *d*=0.82; all other four diagnostic examination tests: *p*>0.22).

To examine the differences between the two patient groups, t-tests were performed for each diagnostic examination time point. Results were summarized in Table 3. No significant differences between the two groups were observed at ‘baseline’ for all five diagnostic tests. Significant differences were observed from the first week of therapy onwards. At diagnostic examination E3, significant differences were observed for the ‘Exploration Task’. At diagnostic examination E4, the Bells-Cancellation-Test, the Copying-Task, and ‘Exploration Test’ differed significantly between the two groups, favoring the *Negami* therapy group. At diagnostic examination E5, this was again the case, now for all neglect tests but the Line-Bisection Task.

**Table 3.** Between-group comparisons for each diagnostic examination time point for all five neglect tests.

|  | Letter-Cancellation | Bells-Cancellation | Copying-Task | Line-Bisection-Task | ‘Exploration<br>Test’ |
| --- | --- | --- | --- | --- | --- |
| Base-<br>line | t(18)=0.4,<br>*p*=0.35 | t(18)=0.47,<br>*p*=0.65 | t(18)=0.21,<br>*p*=0.42 | t(17)=1.58,<br>*p*=0.07 | t(15)=-0.33,<br>*p*=0.75 |
| E3 | t(14)=-1.09,<br>*p*=0.15 | t(13)=-2.02,<br>*p*=0.32 | t(18)=-2.05,<br>*p*=0.03 | t(17)=-0.78,<br>*p*=0.22 | t(13)=-3.9,<br>*p*<0.01*, *d*=25.26 |
| E4 | t(18)=-1.7,<br>*p*=0.05 | t(12)=-2.68,<br>*p*=0.01*,<br>*d*=0.18 | t(18)=-3.64,<br>*p*<0.01*,<br>*d*=1.17 | t(17)=-2.11,<br>*p*=0.03 | t(15)=-5.81,<br>*p*<0.01*, *d*=23 |
| E5 | t(11)=-2.65,<br>*p*=0.01*,<br>*d*=0.18 | t(12)=-3.22,<br>*p*=0.01*,<br>*d*=0.15 | t(18)=-3.84,<br>*p*<0.01*,<br>*d*=1.22 | t(17)=0.46,<br>*p*=0.48 | t(15)=-2.87,<br>*p*<0.01*, *d*=34.6 |

\*= Significant mean differences between groups at the Bonferroni-corrected significance level of 0.0125.

### Follow-up examinations

Pairwise t-comparisons showed no significance for the *Negami* therapy group between time points E4 and E5 for all five neglect tests (all *p*>0.29) as well as between E5 and the follow-up examination E6, again for all neglect tests performed (all *p*>0.35). This indicates that that the improvements due to *Negami* therapy remained stable. Unfortunately, for the ‘Exploration Test’ the latter comparison could not be performed due to missing data at E6 for all subjects. This was caused by an update of the app that was supposed to implement cloud-based data storage (see below discussion). Unfortunately, the update deleted the last collected data, resulting in an irrevocable data loss for the ‘Exploration Test’ of E6. In the group with standard neglect therapy, no significant difference was found between time points E4 and E5 for all five neglect tests (all *p*>0.06).

## Discussion

This study investigated the effectiveness of an AR therapy app for the treatment of spatial neglect after stroke. Two groups, an experimental *Negami* group and a group receiving standard neglect therapy, were compared in a randomized control trial. Both groups were studied in parallel over a five-week period. Both improved significantly under therapy relative to baseline. While the *Negami* therapy group improved in four of the five neglect tests used, the standard therapy group showed improvement only in the Copying-Task. Both these improvements were still seen one week after the end of therapy. The *Negami* therapy group received a second follow-up examination at a time interval between one to two months after completion of training. The improvement remained stable even at this later time point. When we compared the *Negami* therapy group versus the standard neglect therapy group, we found that the improvement of the *Negami* group was significantly superior. The benefit was already evident after one week of therapy (E3) and was also seen after its end. Right after the end of the two weeks training period (E4), the *Negami* therapy group performed better in three neglect tests and one week after the training (E5) in even four of the five neglect tests. Thus, the present results demonstrate not only the efficacy of treating spatial neglect with the *Negami* app but also revealed that the therapy with *Negami* was superior to the standard neglect therapy approach.

From the observation that there was still significant improvement between the first and the second week of *Negami* training, as well as in the standard therapy group, one can deduce that the therapy interval should not be less than at least two weeks. Nevertheless, further studies are pending to assess more precise statements about frequency, extent and duration of therapy.

Effective treatment by exploration training has been shown in several controlled studies, but so far only with small numbers of patients (for review see ^25-28^). An interesting extension of the principle of active exploration training has been the use of virtual reality (VR). Immersive ^29-31^ and non-immersive ^32-34^ VR therapy methods have been tested successfully in (individual) neglect patients. However, the problem of VR methods in general is that a mostly expensive dedicated hardware (e.g., head-mounted display [HMD]) is required ^35^ and that side effects such as dizziness, fatigue, or irritated eyes can occur, termed ‘cybersickness’ ^36,37^. Augmented reality (AR) could be a solution to these problems, since already existing end devices (tablets, cell phones) can be used to run the app and no sensory mismatch is evoked that leads to cybersickness. In fact, when older subjects used the *Negami* app, no or only very minor side effects were observed ^15^.

In the field of AR, another AR-based application for neglect patients has been developed in addition to the *Negami* app. The app asks patients to search for virtual images virtually attached to the walls of the real environment ^38^. While both apps have been shown to increase patient motivation for treatment ^15,38^, the present study is the first that examined the efficacy of such an AR app in treating spatial neglect. Increasing patients’ motivation is especially important considering that motivation for rehabilitation significantly affects clinical outcomes ^39^. This could be one of the reasons why in the present study the *Negami* training group improved significantly more than the patients who received the standard neglect therapy. *Negami* uses various elements of gamification which has been shown to be linked to positive treatment outcome through increased engagement and motivation ^40^.

AR apps are becoming increasingly important also in the treatment of other neurological disorders such as dementia, apraxia, or aphasia. For example, an AR app called “tea making task” has been developed to support patients with apraxia and/or dementia, where the process of making tea has been transformed into a step-by-step instruction system ^41^. The patient receives augmented (virtual) and auditory feedback for each sub-step. Several further AR-based aids have been developed for dementia (for review ^42^). They use virtual texts or images and add them to the patient’s spatial environment by using a wearable AR system in form of a HDM. The idea for this procedure is to use associative learning in order to help the patient to orient and remember better ^42^. AR apps for the treatment of aphasia make use of the real environment to perform augmented naming training, thus ensuring important components of relevance to everyday life ^43^. Early results of testing such augmented and virtual naming training apps suggest efficacy compared to standard treatment, although randomized controlled trials are still lacking ^44,45^.

A big advantage of apps such as *Negami* is that it also can be used after an inpatient stay at a rehabilitation facility in the home setting. In order to maintain and consolidate the positive results after rehabilitation training after discharge, promotion of interventions in the home environment is of crucial importance ^46^. Besides the advantage of maintaining symptom improvement, telerehabilitation could eliminate travel costs to outpatient therapy, or, if patients live in very remote locations, making rehabilitation possible in the first place. On the part of the healthcare system, there may be a reduction in the cost of medical car; asynchronous telerehabilitation allows simultaneous monitoring of several patients by one therapist ^47,48^. *Negami* has the prerequisites to use it for telerehabilitation. By implementing cloud-based synchronization of patient and user management, it ensures that all data collected on the device is automatically synchronized with the cloud when an internet connection is active. This means that the data is then available on all other devices set up by the patient and/or the therapist. This makes working with multiple devices easy. Cloud synchronization is encrypted during data transport to ensure that only authorized persons can access sensitive patient data. Patients can therefore practice from home using the *Negami* app, while therapists and clinicians can access the processed tasks regardless of location to track the patient’s progress and adjust the difficulty level accordingly.

## Conclusion

Two weeks of training with *Negami* in patients with unilateral neglect after stroke significantly improved spatial neglect. Thus, *Negami* can be used as an effective alternative or addition to the current standard neglect therapy and may even be superior to it.

## Data Availability

All data produced in the present study are available upon reasonable request to the authors

## Acknowledgements

This work was supported by Hector Stiftungen, Weinheim (Project M2107).

## Abbreviations

AR: Augmented Reality
CoC: Center of Cancellation
HDM: Head Mounted Display
VR: Virtual Reality

